# A Remote Comprehensive Neurocognitive Test Battery to Monitor Postoperative Neurocognitive Dysfunction in Older Adults: A Prospective Observational Study

**DOI:** 10.64898/2026.02.16.26346401

**Authors:** Mika M. Rockholt, Rachel R. Wu, Bram Seidenberg, Hamleini Martinez, Greta Momesso, Elaine Zhu, Braden V Saba, Raven Perez, Christina Bi, Wonyoung Park, Jessica Hui, Gabrielle Bruno, Daniel Waren, Courtney O’Brien, Romario B. Denoon, Ekow B. Commeh, Vinay K. Aggarwal, Joshua C. Rozell, David Furgiuele, Hyung G. Park, Evan T. Schulze, William Macaulay, Ran Schwarzkopf, Thomas Wisniewski, Ricardo S. Osorio, Lisa V. Doan, Jing Wang

## Abstract

**INTRODUCTION:** Risks for postoperative cognitive dysfunction remain poorly understood. Traditional cognitive screening tools such as the Montreal Cognitive Assessment (MoCA) and the Mini-Mental State Examination (MMSE) are used for perioperative cognitive evaluation but have limited scope, whereas comprehensive in-person testing poses problems for long-term follow up.

**METHODS:** This prospective cohort study assesses the feasibility of using a remotely performed comprehensive neurocognitive test battery, the Uniform Data Set tele-adapted neuropsychological battery version 3 (UDS v3.0 T-cog), administered at baseline and 1 week, 1 month, and 3 months postoperatively, to comprehensively study neurocognitive outcomes in older adults undergoing orthopedic joint arthroplasty. Patient satisfaction with T-cog was assessed through four survey questions evaluating technical issues, duration, willingness to participate in in-person assessment, and satisfaction with remote assessment at 3 months after surgery. Further assessment of pain and mood also included PROMIS scales, McGill Pain Questionnaire, and Pain Catastrophizing Scale, before and 3 months after surgery.

**RESULTS:** 127 participants were enrolled, and out of 120 participants who completed baseline cognitive assessment and underwent surgery, 98 completed cognitive assessments at 3 months. At 3 months, 17% of participants showed an objective decline in cognitive function based on this comprehensive assessment. The remote assessment format was well-received with high participant satisfaction. The UDS v3.0 T-cog identified deficits in specific domains that would have been missed by brief screening instruments, supporting its values for perioperative use.

**DISCUSSION:** This is the first study to utilize this comprehensive remote cognitive assessment tool to study long-term cognitive function. The assessment can be combined with other preoperative outcome assessments in older adults undergoing surgery.

**Highlights:**

- Current detection of perioperative cognitive outcomes in older adults rely on in-person cognitive assessments that are varied in methodology and often lack sensitivity and specificity
- The UDS v3.0 T-cog identified objective cognitive decline in 17% of patients after orthopedic arthroplasty while also detecting early non-memory cognitive decline through the more comprehensive test battery with high participant satisfaction and retention, supporting remote assessment feasibility.
- These findings suggest that remote comprehensive cognitive assessments are an effective tool to provide early detection and risk stratification for perioperative neurocognitive dysfunction in older patients.

## 1 BACKGROUND

Historically, postoperative cognitive dysfunction (POCD) describes an objective decline in cognitive function detected one week or more after surgery ^1^, and it has been reported to occur in 2.2–50% of non-cardiac surgeries ^2–11^. More recently, the term perioperative neurocognitive disorders (NCDs) has been adopted to indicate cognitive decline that can be both short- and long-term. NCDs negatively impact recovery, diminish long-term quality of life, and increase both morbidity and mortality ^7, 12, 13^, with advanced age, chronic pain, and preexisting cognitive impairment—especially mild cognitive impairment (MCI), affecting 10–20% of older adults—being key risk factors ^4, 7^. This highlights the importance of early identification and risk stratification ^2, 7, 14, 15^.

A key barrier in research on NCDs has been the variation, specificity, and sensitivity of the tests administered ^6, 16^. Common tools like the Montreal Cognitive Assessment (MoCA) and the Mini-Mental State Examination (MMSE) do not adequately capture multiple cognitive domains and may lack sensitivity, prompting interest in more extensive neuropsychological batteries that assess multiple cognitive domains ^2, 3, 7, 10, 16, 17^. Another key challenge in NCD research is the difficulty in maintaining long-term follow-up ^9, 18^, as comprehensive in-person neuropsychological testing can pose substantial burden in older adults whose mobility and independence may be compromised after surgery ^18^. The Uniform Data Set tele-adapted cognitive battery (UDS v3.0 T-Cog), developed by the National Alzheimer’s Coordinating Center ^19, 20^, can address these barriers by enabling remote, serial cognitive assessments capturing multiple cognitive domains. Despite widespread use in cognitive research, it is rarely applied in perioperative contexts.

Chronic pain affects over 50% of adults aged 65 and older, with osteoarthritis—particularly of the hip and knee—being a leading contributor to pain and functional decline ^21^. As the U.S. population ages, orthopedic surgeries have increased, now accounting for a considerable proportion of procedures performed in this age group ^22^. Hip and knee arthroplasties significantly improve osteoarthritic symptoms and functional status in older adults ^22^. However, despite clinical benefits, concerns around postoperative complications, including NCDs are growing ^1, 23^. POCD has been reported to occur in 10–45% of orthopedic surgeries ^2–11^, and the large range of incidence reflects variations in study methods, sensitivity and specificity. Interestingly, most studies report no significant difference in POCD incidence when comparing general versus regional anesthesia ^24, 25^.

Here, we assessed the feasibility of using the T-Cog battery to define the incidence of NCD following total hip and knee arthroplasty in older adults. Given the comorbidity of chronic pain in this population, we also assessed whether cognitive testing could be feasibly integrated with comprehensive pain, mood, and sleep evaluations. To our knowledge, it is the first study to combine comprehensive remote cognitive testing and perioperative pain and other behavioral evaluation in an older total joint arthroplasty population.

## 2 METHODS

### 2.1 Study participants

Data for this longitudinal cohort study were obtained from an ongoing, prospective, cohort study of patients undergoing elective total knee arthroplasty (TKA) or total hip arthroplasty (THA). We enrolled a total of 127 patients recruited from a tertiary academic medical center from November 25, 2023, to May 27, 2025 (**Figure 1**). Eligible participants were aged ≥ 65 years, English-speaking and able to communicate verbally, with American Society of Anesthesiologist physical status of ≤ III. Exclusion criteria were pre-existing dementia, history of schizophrenia, epilepsy, craniotomy or cerebrovascular accident (stroke and/or hemorrhage), failure to pass a hearing verification before beginning the remote cognitive assessment, and unwillingness to give informed consent.

**Figure 1.**
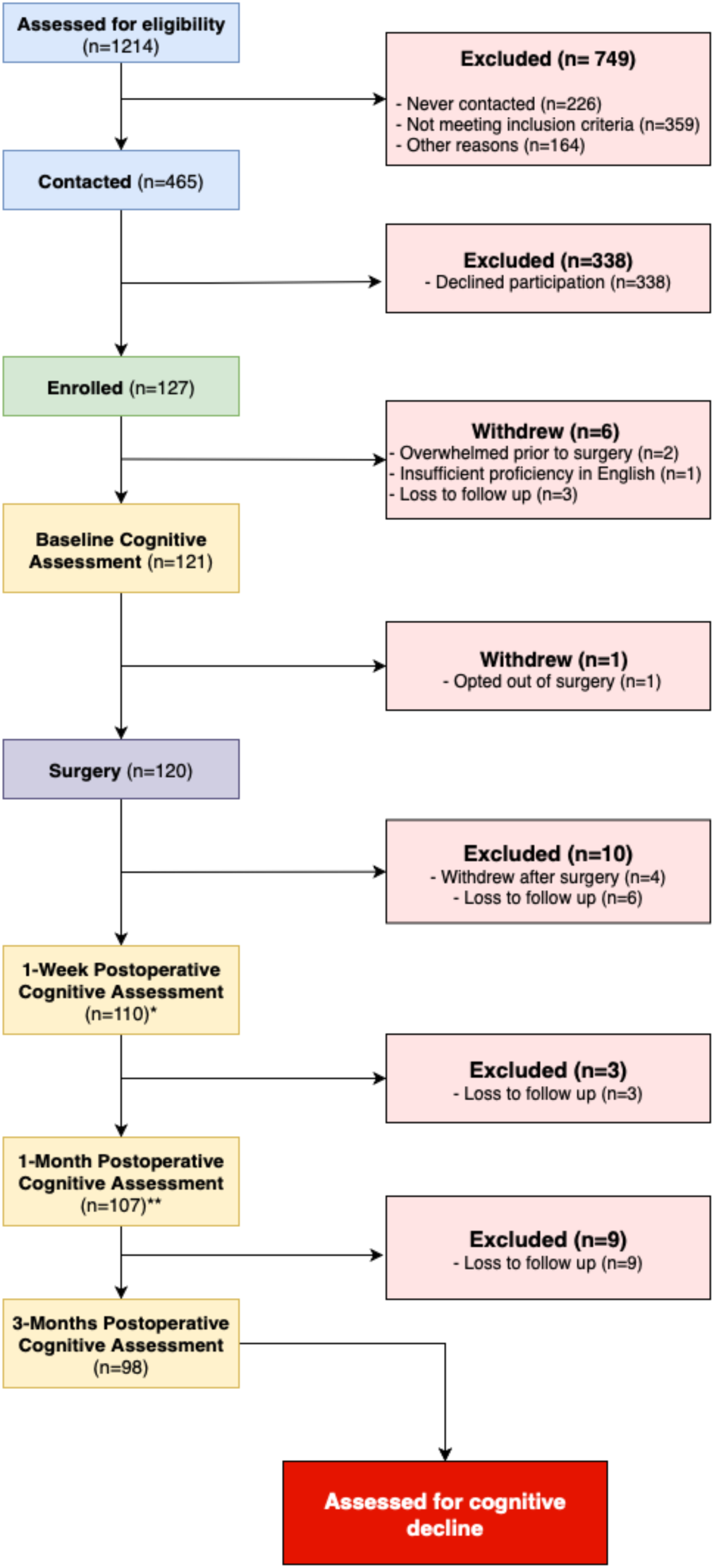
STROBE flow diagram showing the screening, enrollment, and selection process of participants assessed for cognitive decline. Of the 1214 individuals screened for eligibility, 465 (38%) eligible candidates were contacted. Among those contacted, 338 individuals declined participation. The primary reasons for non-participation included a lack of interest in research involvement (56%), time constraints or limited availability (31%), and feeling overwhelmed by surgical demands and the postoperative recovery period (10%). 127 participants were enrolled in the study, corresponding to an enrollment rate of 27%. 120 participants completed the baseline assessment and underwent surgery. Of the 107 participants who completed the 1-month follow-up assessment, 9 were lost to follow-up at the 3-month mark. As of the date of writing, 98 out of 120 participants who underwent surgery have completed the 3-month cognitive assessment, yielding a retention rate of 82%. Figure was created in BioRender (www.biorender.com). ***Flowchart depicting selection of participants*** *\* A total of 98 subjects (89.1%) finished their 1-week postoperative cognitive assessment, 12 subjects (10.9%) cancelled due to feeling unwell, due to hospitalization, or forgetting their appointment and re-joined for the following assessment*. *\*\*A total of 99 subjects (92.5%) completed the 1 month assessment, while 8 subjects (7.5%) cancelled due to feeling unwell, due to hospitalization, or forgetting their appointment but (unless they were lost to follow up) re-joined for the following assessment*.

### 2.2 Data collection

#### Demographic data and medical history

Patient age, medical and medication history were extracted from electronic health records (Epic Systems Corporation, Madison, WI, USA). Patient-reported data on sex, race, ethnicity, and highest level of education were captured via Research Electronic Data Capture (REDCap) HIPAA-compliant online surveys.

#### Objective assessment of cognitive function

All cognitive assessments were conducted remotely using the National Alzheimer’s Coordinating Center’s (NACC) tele-adapted protocols ^19, 20^. The Uniform Data Set (UDS) Version 3.0 tele-cognitive battery (v3.0 T-cog; Form C2T, May 2020) consists of ten neuropsychological tests designed to evaluate a range of cognitive domains including: global functioning through the MoCA Blind (used to assess baseline cognitive impairment but was not included in the determination of objective cognitive decline at three months), executive function/processing speed through the Number Span Test (Backward) and the Oral Trail Making Test B, learning/memory through the Craft Story 21 Recall and the Rey Auditory Verbal Learning Test (RAVLT), attention/processing speed through the Number Span Test (Forward) and the Oral Trail Making Test A, and language through the Category Fluency Test (Animals and Vegetables for semantic language), the Verbal Fluency Test with F and L words (for phonemic language), and the Verbal Naming Test (for auditory responsive naming) (**Supplemental Figure 1**) ^20^. The battery was administered according to participant preference, either via telephone or a HIPAA-compliant video conferencing platform (Zoom; Zoom Video Communications Inc, San Jose, CA, USA) at baseline and 1 week, 1 month, and 3 months postoperatively. Participants were instructed to complete the assessments independently, without assistance, in a quiet setting, free from distractions, and with a stable telephone or internet connection. With consent, the sessions were audio recorded to ensure quality assurance in staff training and to verify participant responses. Cognitive evaluations were performed by a trained research team member under the supervision of a licensed neuropsychologist. The UDS v3.0 T-cog includes a validity checklist for researchers to document factors potentially affecting test performance, such as environmental distractions, participant disengagement, or hearing impairments. At the conclusion of each assessment, research team members rated the validity of results as “Very valid,” “Questionably valid,” or “Invalid,” providing context on testing conditions.

#### Subjective assessment of cognitive function

To evaluate subjective cognitive complaints (SCC), participants were asked to answer an open-ended question (yes with explanation in free text/no) and/or the 20-item cognitive change index (CCI-20) – a widely used tool to assess SCC ^26^– to describe whether they noticed any cognitive changes 3 months after their surgery. The CCI-20 responds to items on a 1 to 5 Likert scale with higher scores indicating greater decline (1 = no change or normal ability, 2 = minimal change or slight/occasional problem, 3 = some change or mild problem, 4 = clearly noticeable change or moderate problem, 5 = much worse or severe problem) ^26^. The CCI-20 was added during the course of the study as a validated measure of SCC ^26^.

#### Comprehensive pain assessment

Pain, mood, and sleep were assessed using validated questionnaires, including the Patient Reported Outcomes Measurement Information System (PROMIS) measures (Pain Intensity 3a, Neuropathic Pain 5a, Pain Interference 6a, Anxiety 4a, Depression 4a, Physical Function 4a, and Sleep Disturbance 4a) ^27^, the McGill Pain Questionnaire (MPQ) ^28^, and the Pain Catastrophizing Scale (PCS) ^29^. PROMIS results are reported as average T-scores, standardized according to established scoring guidelines (https://www.healthmeasures.net/) ^27^. Except for the Physical Function 4a scale, higher PROMIS T-scores indicate greater symptom severity. The MPQ evaluates the qualitative aspects of pain, with higher scores reflecting increased pain experience^28^. The PCS quantifies cognitive and emotional responses to pain; elevated scores denote greater pain catastrophizing ^29^. Pain, mood, and sleep questionnaires were distributed prior to surgery and again at three months postoperatively.

#### Assessment of patient satisfaction

Participant satisfaction was assessed through four survey questions evaluating technical accessibility, assessment duration, willingness to participate in in-person assessments, and overall satisfaction with the remote cognitive assessment ^20^.

### 2.3 Statistical Analyses

#### Pre-existing cognitive impairment

Participants with baseline scores below 18 out of 22 on the MoCA Blind component of the UDS v3.0 T-Cog battery were classified as having pre-existing cognitive impairment, a term referring to participants with objectively measurable cognitive impairment at baseline ^1^.

#### Postoperative neurocognitive disorders

Z-scores for most tests were calculated using a regression-based, normative score calculator ^30^. The RAVLT Z-scores were derived from the Mayo Normative Studies Scoring Resource, which used data from the Mayo Older Americans Normative Studies (MOANS; ^31^). Oral Trail Making test scores were calculated using normative data from Mrazik et al. ^32^, as recommended by the NACC. Objective criteria for a decline in cognitive function was defined as a decline of ≥1 standard deviation in normative based Z-scores—relative to the individual’s own baseline—observed at the postoperative time point on at least two cognitive tests in the UDS v3.0 T-cog test battery within the same domain ^1, 6, 9, 16, 23^. Requiring at least two tests within the same domain reduces the likelihood that test performance was influenced by chance or temporary inattention.

Continuous variables were presented as mean (± standard deviation or min-max), and categorical variables as a number (percentage). Long-term follow-up retention rates were calculated based on all enrolled subjects remaining in the study who completed their 3-month assessment within the designated 3-month time frame. Subjects with pending 3-month assessments were excluded from the final retention rate calculation. Exploratory testing of associations between baseline pain/mood measures and 3-month cognitive outcomes was conducted by comparing the group of patients who did not demonstrate cognitive decline at 3-months with the group who did, using the chi-square test for categorical variables and the non-parametric Mann-Whitney *U*-test and/or unpaired t-test for continuous variables. Likewise, exploratory testing of associations between 3-month pain/mood measures and 3-month cognitive outcomes was conducted with the same methods. For multiple comparisons, significance was adjusted using the Bonferroni correction. A two-tailed P-value <0.05 was considered statistically significant. To identify independent risk factors for cognitive decline at three months, baseline data - including pre-existing cognitive impairment, scores from the UDS v3.0 T-cog test battery, and behavioral data from pain, mood, and sleep questionnaires —were analyzed using univariate logistic regressions. The Hosmer-Lemeshow test for the goodness of fit was used for univariate testing. No multivariate logistic regressions were performed due to the low number of outcomes in the cognitive decline group (n = 17). Statistical analyses were conducted using Microsoft Excel version 16.91 (Microsoft Corporation, Redmond, WA, USA), GraphPad Prism version 10.3.1 (GraphPad Software, Boston, MA, USA), and IBM SPSS 30 (IBM Corp. Armonk, NY, USA).

## 3 RESULTS

One hundred twenty-one participants underwent baseline cognitive assessment, of whom 120 underwent surgery. At 3-month follow-up, a total of 98 patients had completed the cognitive assessment and were therefore included in the analysis (**Figure 1**). Of these, 42 were male (43%) and 56 were female (57%), with a mean age of 73.69 ± 5.2 years. The most prevalent comorbidities were hypertension (67%), hypercholesterolemia (66%), and obesity (37%). A total of 56% of patients underwent total knee arthroplasty, while 44% underwent total hip arthroplasty. The majority of procedures (95%) were performed under regional anesthesia. Baseline characteristics are detailed in **Table 1**.

**Table 1.** Baseline characteristics.

| Demographics | Cognitive Decline at 3-Months |  | p-value |
| --- | --- | --- | --- |
|  | No (n = 81) | Yes (n = 17) |  |
| **Gender, (%)** |  |  | 0.023 |
| - Male | 39 (48.1) | 3 (17.6) |  |
| - Female | 42 (51.9) | 14 (82.4) |  |
| **Age, mean (± SD)** | 73.6 ± 5.1 | 74.1 ± 6.0 | 0.829 |
| **Age Group, range (%)** |  |  | 0.357 |
| - 65-69 | 21 (25.9) | 4 (23.5) |  |
| - 70-74 | 18 (22.2) | 5 (29.4) |  |
| - 75-79 | 35 (43.2) | 5 (29.4) |  |
| - 80-84 | 5 (6.2) | 1 (5.9) |  |
| - 85 + | 2 (2.5) | 2 (11.8) |  |
| **Years of Education, mean (± SD)<sup>1</sup>** | 17.5 ± 3.0 | 18 ± 2.7 | 0.560 |
| **Level of Education** |  |  | 0.232 |
| - High school diploma | 7 (8.6) | 1 (5.9) |  |
| - Associate degree | 13 (16.0) | 1 (5.9) |  |
| - Bachelor's degree | 16 (19.8) | 4 (23.5) |  |
| - Doctoral degree | 44 (54.3) | 9 (52.9) |  |
| - Missing Data | 1 (1.2) | 2 (11.8) |  |
| **Race (%)** |  |  | 0.041 |
| - White | 65 (80.2) | 10 (58.8) |  |
| - Black/African American | 12 (14.8) | 3 (17.6) |  |
| - Asian | 1 (1.2) | 2 (11.8) |  |
| - American Indian/Alaska Native | 0 (0.0) | 0 (0.0) |  |
| - Unknown<sup>2</sup> | 3 (3.7) | 2 (11.8) |  |
| **Ethnicity (%)** |  |  | 0.212 |
| - Hispanic or Latino | 9 (11.1) | 1 (5.9) |  |
| - Not Hispanic or Latino | 65 (80.2) | 12 (70.6) |  |
| - Not Reported/Unknown<sup>2</sup> | 7 (8.6) | 4 (23.5) |  |
| **Past Medical History, yes (%)** |  |  |  |
| - Hypertension | 54 (66.7) | 12 (70.6) | 0.754 |
| - Hypercholesterolemia and/or hyperlipidemia | 56 (69.1) | 9 (52.9) | 0.199 |
| - Obesity | 27 (33.3) | 9 (52.9) | 0.127 |
| - History of Malignancy (Active or Past) | 27 (33.3) | 4 (23.5) | 0.429 |
| - Thyroid Disease | 17 (21.0) | 5 (29.4) | 0.449 |
| - Anxiety | 9 (11.1) | 3 (17.6) | 0.432 |
| - Depression | 12 (14.8) | 4 (23.5) | 0.469 |
| - Lung Disease (Asthma and/or COPD) | 8 (9.9) | 2 (11.8) | 0.683 |
| - Coronary Artery Disease | 14 (17.3) | 3 (17.6) | 0.971 |
| - Diabetes | 17 (21.0) | 3 (17.6) | 0.756 |
| - Sleep Apnea | 13 (16.0) | 4 (23.5) | 0.487 |
| **Smoking History (Current or Former), yes (%)** | 39 (48.1) | 9 (52.9) | 0.719 |
| **History of Alcohol Overconsumption, yes (%)** | 13 (16.0) | 1 (5.9) | 0.453 |
| **Medication History (%)** |  |  |  |
| - Opioids | 24 (29.6) | 6 (35.3) | 0.645 |
| - Anti-inflammatory Agents (NSAIDs) | 42 (51.9) | 9 (52.9) | 0.935 |
| - Gabapentinoids | 9 (11.1) | 2 (11.8) | 0.938 |
| - Benzodiazepines | 7 (8.6) | 2 (11.8) | 0.653 |
| - Antidepressants | 10 (12.3) | 5 (29.4) | 0.130 |
| **Type of Surgery, (%)** |  |  | 0.433 |
| - Total Knee Arthroplasty | 44 (54.3) | 11 (64.7) |  |
| - Total Hip Arthroplasty | 37 (45.7) | 6 (35.3) |  |
| **Type of Anesthesia, (%)** |  |  | 0.872 |
| - Regional Anesthesia | 77 (95.1) | 16 (94.1) |  |
| - General Anesthesia | 4 (4.9) | 1 (5.9) |  |
| **Cognitive Impairment at Baseline** | 17 (21.0) | 6 (35.3) | 0.219 |

<sup>1</sup> Years of education were totaled based on completed degrees: high school = 12 years, associate degree = 2 years bachelors = 4 years, masters = 2 years, doctoral = 4 years
<sup>2</sup> Patients preferred not to respond

### 3.1 Cognitive Outcomes

At baseline, 23 of 98 participants who completed 3-month cognitive assessments (24%) exhibited pre-existing cognitive impairment. Among them, 5 (22%) had an objective decline in cognitive function at three months. Overall, 17 of these 98 participants (17%) had an objective decline in cognitive function at the 3-month follow-up. These individuals demonstrated significantly lower performance in processing speed, executive function, attention, and language at baseline (p < 0.05). At the 3-month follow-up, they also showed significantly lower scores in several assessments within the learning/memory and language domains. In contrast, participants who did not exhibit objective decline in cognitive function at three months showed significant improvements across all tests in the learning and memory domain, as well as some tests in the executive function and language domains. Despite significant improvements in most pain, mood and sleep measures, there were no differences in cognitive outcomes when comparing between types of surgery (**Supplemental Table 1**). Detailed cognitive performance results are presented in **Table 2**, with a subgroup analysis presented in **Supplemental Table 2.**

**Table 2.**
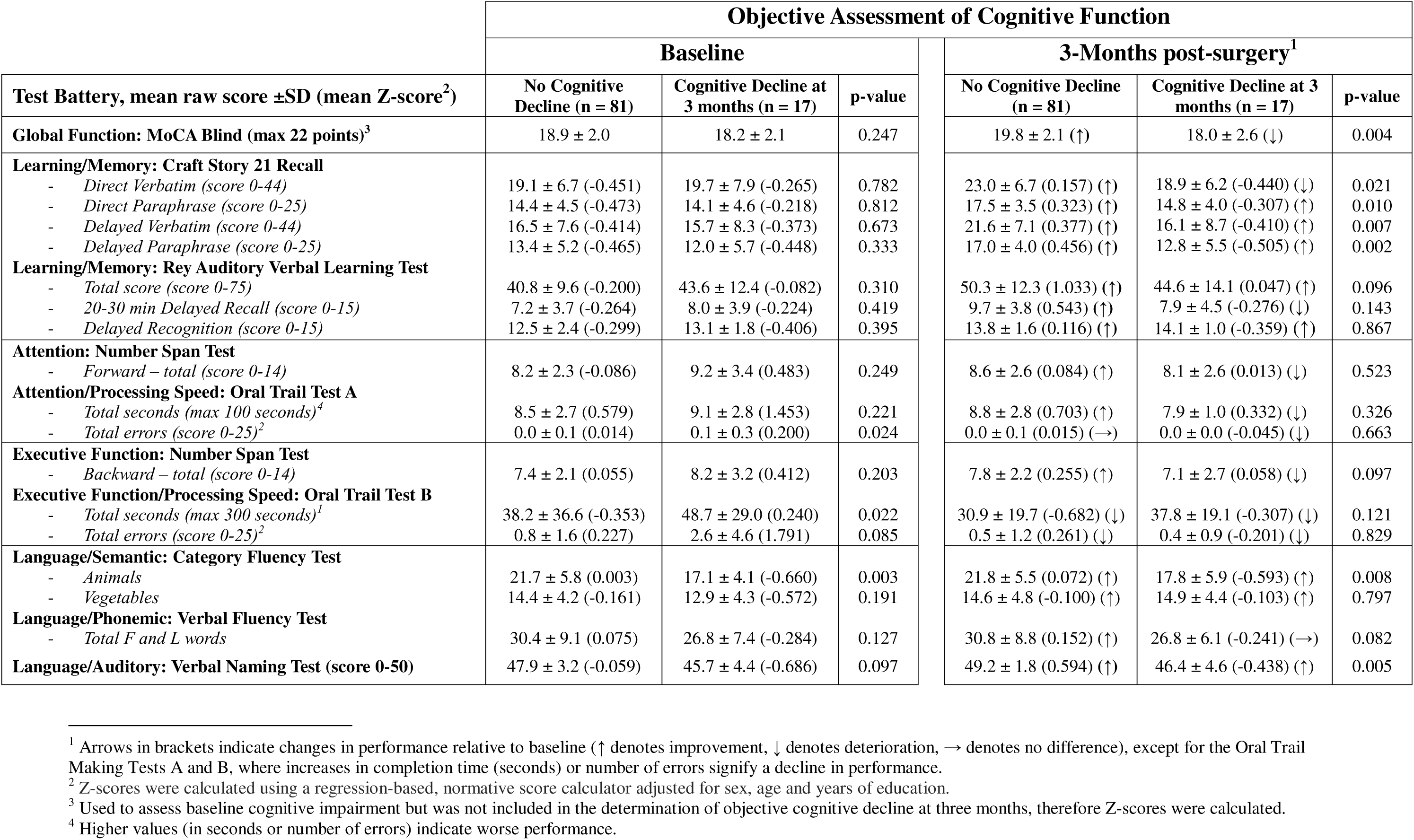
Objective assessment of cognitive function.

| Test Battery, mean raw score ±SD (mean Z-score<sup>2</sup>) | Objective Assessment of Cognitive Function | | | | | |
| --- | --- | --- | --- | --- | --- | --- |
|  | Baseline |  |  | 3-Months post-surgery<sup>1</sup> |  |  |
|  | No Cognitive Decline (n = 81) | Cognitive Decline at 3 months (n = 17) | p-value | No Cognitive Decline (n = 81) | Cognitive Decline at 3 months (n = 17) | p-value |
| **Global Function: MoCA Blind (max 22 points)<sup>3</sup>** | 18.9 ± 2.0 | 18.2 ± 2.1 | 0.247 | 19.8 ± 2.1 (↑) | 18.0 ± 2.6 (↓) | 0.004 |
| **Learning/Memory: Craft Story 21 Recall** |  |  |  |  |  |  |
| - Direct Verbatim (score 0-44) | 19.1 ± 6.7 (-0.451) | 19.7 ± 7.9 (-0.265) | 0.782 | 23.0 ± 6.7 (0.157) (↑) | 18.9 ± 6.2 (-0.440) (↓) | 0.021 |
| - Direct Paraphrase (score 0-25) | 14.4 ± 4.5 (-0.473) | 14.1 ± 4.6 (-0.218) | 0.812 | 17.5 ± 3.5 (0.323) (↑) | 14.8 ± 4.0 (-0.307) (↑) | 0.010 |
| - Delayed Verbatim (score 0-44) | 16.5 ± 7.6 (-0.414) | 15.7 ± 8.3 (-0.373) | 0.673 | 21.6 ± 7.1 (0.377) (↑) | 16.1 ± 8.7 (-0.410) (↑) | 0.007 |
| - Delayed Paraphrase (score 0-25) | 13.4 ± 5.2 (-0.465) | 12.0 ± 5.7 (-0.448) | 0.333 | 17.0 ± 4.0 (0.456) (↑) | 12.8 ± 5.5 (-0.505) (↑) | 0.002 |
| **Learning/Memory: Rey Auditory Verbal Learning Test** |  |  |  |  |  |  |
| - Total score (score 0-75) | 40.8 ± 9.6 (-0.200) | 43.6 ± 12.4 (-0.082) | 0.310 | 50.3 ± 12.3 (1.033) (↑) | 44.6 ± 14.1 (0.047) (↑) | 0.096 |
| - 20-30 min Delayed Recall (score 0-15) | 7.2 ± 3.7 (-0.264) | 8.0 ± 3.9 (-0.224) | 0.419 | 9.7 ± 3.8 (0.543) (↑) | 7.9 ± 4.5 (-0.276) (↓) | 0.143 |
| - Delayed Recognition (score 0-15) | 12.5 ± 2.4 (-0.299) | 13.1 ± 1.8 (-0.406) | 0.395 | 13.8 ± 1.6 (0.116) (↑) | 14.1 ± 1.0 (-0.359) (↑) | 0.867 |
| **Attention: Number Span Test** |  |  |  |  |  |  |
| - Forward – total (score 0-14) | 8.2 ± 2.3 (-0.086) | 9.2 ± 3.4 (0.483) | 0.249 | 8.6 ± 2.6 (0.084) (↑) | 8.1 ± 2.6 (0.013) (↓) | 0.523 |
| **Attention/Processing Speed: Oral Trail Test A** |  |  |  |  |  |  |
| - Total seconds (max 100 seconds)<sup>4</sup> | 8.5 ± 2.7 (0.579) | 9.1 ± 2.8 (1.453) | 0.221 | 8.8 ± 2.8 (0.703) (↑) | 7.9 ± 1.0 (0.332) (↓) | 0.326 |
| - Total errors (score 0-25)<sup>2</sup> | 0.0 ± 0.1 (0.014) | 0.1 ± 0.3 (0.200) | 0.024 | 0.0 ± 0.1 (0.015) (→) | 0.0 ± 0.0 (-0.045) (↓) | 0.663 |
| **Executive Function: Number Span Test** |  |  |  |  |  |  |
| - Backward – total (score 0-14) | 7.4 ± 2.1 (0.055) | 8.2 ± 3.2 (0.412) | 0.203 | 7.8 ± 2.2 (0.255) (↑) | 7.1 ± 2.7 (0.058) (↓) | 0.097 |
| **Executive Function/Processing Speed: Oral Trail Test B** |  |  |  |  |  |  |
| - Total seconds (max 300 seconds)<sup>1</sup> | 38.2 ± 36.6 (-0.353) | 48.7 ± 29.0 (0.240) | 0.022 | 30.9 ± 19.7 (-0.682) (↓) | 37.8 ± 19.1 (-0.307) (↓) | 0.121 |
| - Total errors (score 0-25)<sup>2</sup> | 0.8 ± 1.6 (0.227) | 2.6 ± 4.6 (1.791) | 0.085 | 0.5 ± 1.2 (0.261) (↓) | 0.4 ± 0.9 (-0.201) (↓) | 0.829 |
| **Language/Semantic: Category Fluency Test** |  |  |  |  |  |  |
| - Animals | 21.7 ± 5.8 (0.003) | 17.1 ± 4.1 (-0.660) | 0.003 | 21.8 ± 5.5 (0.072) (↑) | 17.8 ± 5.9 (-0.593) (↑) | 0.008 |
| - Vegetables | 14.4 ± 4.2 (-0.161) | 12.9 ± 4.3 (-0.572) | 0.191 | 14.6 ± 4.8 (-0.100) (↑) | 14.9 ± 4.4 (-0.103) (↑) | 0.797 |
| **Language/Phonemic: Verbal Fluency Test** |  |  |  |  |  |  |
| - Total F and L words | 30.4 ± 9.1 (0.075) | 26.8 ± 7.4 (-0.284) | 0.127 | 30.8 ± 8.8 (0.152) (↑) | 26.8 ± 6.1 (-0.241) (→) | 0.082 |
| **Language/Auditory: Verbal Naming Test (score 0-50)** | 47.9 ± 3.2 (-0.059) | 45.7 ± 4.4 (-0.686) | 0.097 | 49.2 ± 1.8 (0.594) (↑) | 46.4 ± 4.6 (-0.438) (↑) | 0.005 |

<sup>1</sup> Arrows in brackets indicate changes in performance relative to baseline ( ↑ denotes improvement, ↓ denotes deterioration, → denotes no difference), except for the Oral Trail Making Tests A and B, where increases in completion time (seconds) or number of errors signify a decline in performance.
<sup>2</sup> Z-scores were calculated using a regression-based, normative score calculator adjusted for sex, age and years of education.
<sup>3</sup> Used to assess baseline cognitive impairment but was not included in the determination of objective cognitive decline at three months, therefore Z-scores were calculated.
<sup>4</sup> Higher values (in seconds or number of errors) indicate worse performance.

Out of 81 subjects who filled in the questions regarding SCC, 30 subjects reported SCC (37%; 30/81). Of those, 5 subjects showed objective cognitive decline at 3 months (**Supplemental Table 2**). Of those who self-reported CI at 3 months but did not have enough measured decline on cognitive testing to be classified with POCD at 3 months (n = 24, 24/81 = 30%), one had a decline in executive functioning and processing speed at 1 week, two had declines in learning and memory at 1 week, while the rest had no decline at 1 week, 1 month or 3 months.

### 3.2 Feasibility and Acceptability of Remote Cognitive Assessments

The feasibility of the UDS v3.0 T-cog was assessed at multiple time points: baseline, 1 week, 1 month and 3 months after surgery. These time points were chosen due to their relevance in perioperative cognitive research. Baseline is needed to understand a change in cognitive performance. One week and 1 month time points are chosen to reflect subacute cognitive changes after surgery. The 3-month time point is used for assessing long-term cognitive changes. A total of 416 remote cognitive assessments were completed across the study timeline: 121 at baseline, 98 at 1 week, 99 at 1 month, and 98 at 3 months. Of these, 308 assessments (74%) were conducted via videoconferencing. Specifically, 89 assessments (74%) at baseline, 72 (73%) at 1 week, 77 (78%) at 1 month, and 70 (71%) at 3 months were video based. The mean duration of each assessment was 51.2 minutes (range: 40–70 minutes). Assessments classified as “questionably valid” were attributed to distractions (35%), lack of effort (25%), interruptions (10%), and fatigue (10%). Among participants who had objective decline in cognitive function at three months, one had a questionably valid assessment at one month due to lack of effort or disinterest. At the 3-month mark, four participants with objective decline in cognitive function had questionably valid assessments, primarily due to distractions (3/4, 75%) and one due to potential language barrier. (**Supplemental Table 2**).

Overall, most patients found the remote cognitive assessments technically very easy (43%) or easy (35%) to complete and appropriately timed in duration (52%). Additionally, 50% indicated they would be less likely to participate if the assessments were conducted in person. The remote assessment protocol achieved a 99% satisfaction rate (very satisfied: 48%, satisfied: 51%) (**Supplemental Figure 2**)

### 3.3 Preoperative Pain, Mood and Sleep Assessment

The results on pain, mood and sleep questionnaires are presented in **Table 3**. Prior to surgery, there were no significant differences in baseline questionnaires between patients who developed objective decline in cognitive function and those who did not. At 3 months, there were also no significant differences in the scores overall between the two groups.

**Table 3.**
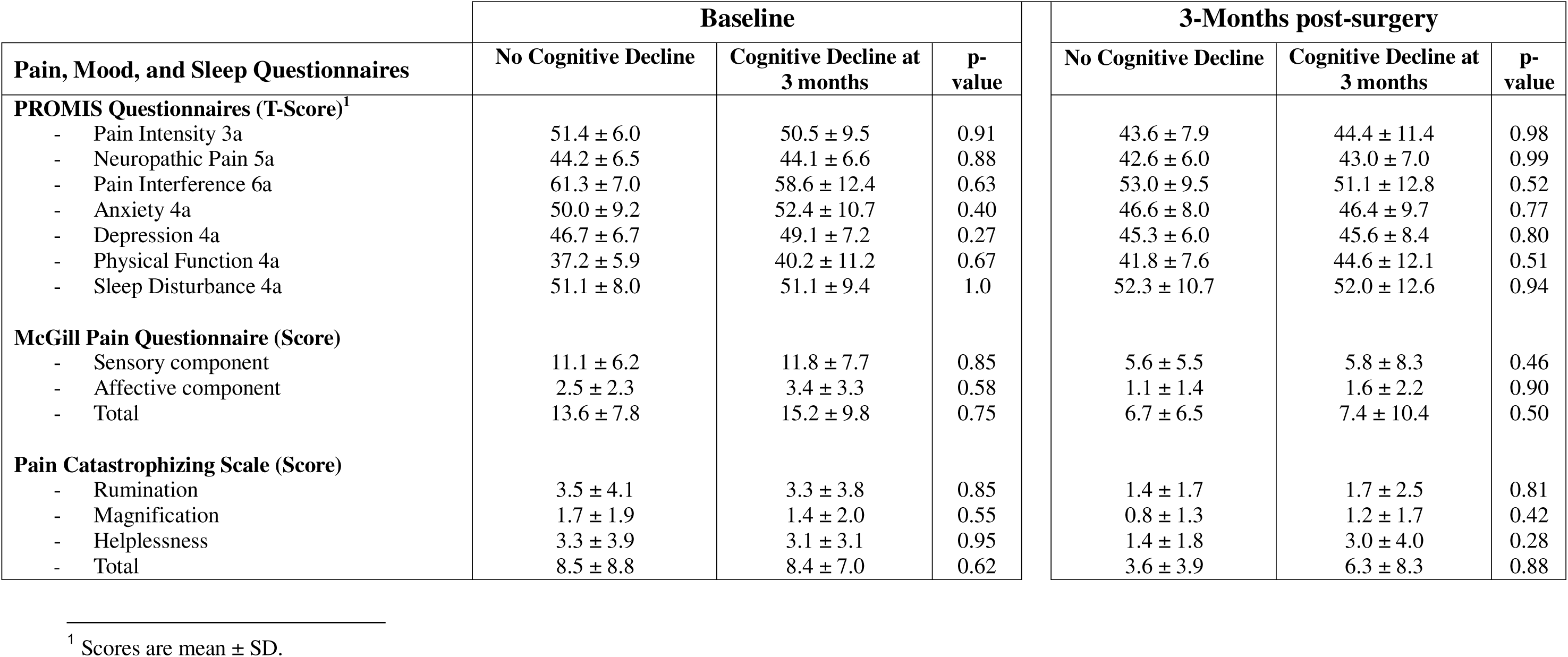
Perioperative pain, mood and sleep assessments in our patient cohort.

| Pain, Mood, and Sleep Questionnaires | Baseline |  |  | 3-Months post-surgery |  |  |
| --- | --- | --- | --- | --- | --- | --- |
|  | No Cognitive Decline | Cognitive Decline at 3 months | p-value | No Cognitive Decline | Cognitive Decline at 3 months | p-value |
| **PROMIS Questionnaires (T-Score)<sup>1</sup>** |  |  |  |  |  |  |
| - Pain Intensity 3a | 51.4 ± 6.0 | 50.5 ± 9.5 | 0.91 | 43.6 ± 7.9 | 44.4 ± 11.4 | 0.98 |
| - Neuropathic Pain 5a | 44.2 ± 6.5 | 44.1 ± 6.6 | 0.88 | 42.6 ± 6.0 | 43.0 ± 7.0 | 0.99 |
| - Pain Interference 6a | 61.3 ± 7.0 | 58.6 ± 12.4 | 0.63 | 53.0 ± 9.5 | 51.1 ± 12.8 | 0.52 |
| - Anxiety 4a | 50.0 ± 9.2 | 52.4 ± 10.7 | 0.40 | 46.6 ± 8.0 | 46.4 ± 9.7 | 0.77 |
| - Depression 4a | 46.7 ± 6.7 | 49.1 ± 7.2 | 0.27 | 45.3 ± 6.0 | 45.6 ± 8.4 | 0.80 |
| - Physical Function 4a | 37.2 ± 5.9 | 40.2 ± 11.2 | 0.67 | 41.8 ± 7.6 | 44.6 ± 12.1 | 0.51 |
| - Sleep Disturbance 4a | 51.1 ± 8.0 | 51.1 ± 9.4 | 1.0 | 52.3 ± 10.7 | 52.0 ± 12.6 | 0.94 |
| **McGill Pain Questionnaire (Score)** |  |  |  |  |  |  |
| - Sensory component | 11.1 ± 6.2 | 11.8 ± 7.7 | 0.85 | 5.6 ± 5.5 | 5.8 ± 8.3 | 0.46 |
| - Affective component | 2.5 ± 2.3 | 3.4 ± 3.3 | 0.58 | 1.1 ± 1.4 | 1.6 ± 2.2 | 0.90 |
| - Total | 13.6 ± 7.8 | 15.2 ± 9.8 | 0.75 | 6.7 ± 6.5 | 7.4 ± 10.4 | 0.50 |
| **Pain Catastrophizing Scale (Score)** |  |  |  |  |  |  |
| - Rumination | 3.5 ± 4.1 | 3.3 ± 3.8 | 0.85 | 1.4 ± 1.7 | 1.7 ± 2.5 | 0.81 |
| - Magnification | 1.7 ± 1.9 | 1.4 ± 2.0 | 0.55 | 0.8 ± 1.3 | 1.2 ± 1.7 | 0.42 |
| - Helplessness | 3.3 ± 3.9 | 3.1 ± 3.1 | 0.95 | 1.4 ± 1.8 | 3.0 ± 4.0 | 0.28 |
| - Total | 8.5 ± 8.8 | 8.4 ± 7.0 | 0.62 | 3.6 ± 3.9 | 6.3 ± 8.3 | 0.88 |

<sup>1</sup> Scores are mean ± SD.

Previously described risk factors for perioperative cognitive disorders were chosen as independent variables to assess potential associations with the development of objective decline in cognitive function at 3 months. In the univariate logistic regression analysis, presented in **Table 4**, there were no definitive associations with objective decline in cognitive function at 3 months. However, this study is not powered for comprehensive analysis of the association between pre- and postoperative mood and pain outcomes and the development of postoperative cognitive decline.

**Table 4.** Univariate regression analysis.

| Independent variable<sup>2</sup> | pNCD at 3 months |  | Univariate Analyses<sup>1</sup> |  |  |
| --- | --- | --- | --- | --- | --- |
|  | No<br>n = 81 | Yes<br>n = 17 | OR | 95% CI | p-value |
| Age | - | - | 1.019 | 0.921 – 1.127 | 0.717 |
| Years of Education | - | - | 1.063 | 0.872 – 1.296 | 0.545 |
| Past Medical History |  |  |  |  |  |
| - Hypertension | 54 (66.7) | 12 (70.6) | 1.200 | 0.383 – 3.756 | 0.754 |
| - Obesity | 27 (33.3) | 9 (52.9) | 2.250 | 0.781 – 6.485 | 0.133 |
| - Diabetes | 17 (21.0) | 3 (17.6) | 0.807 | 0.208 – 3.133 | 0.756 |
| - Hypercholesterolemia/Hyperlipidemia | 56 (69.1) | 9 (52.9) | 0.502 | 0.174 – 1.454 | 0.204 |
| Smoking History | 39 (48.1) | 9 (52.9) | 1.212 | 0.425 – 3.453 | 0.720 |
| Medication History |  |  |  |  |  |
| - Antidepressants | 10 (12.3) | 5 (29.4) | 2.958 | 0.860 – 10.178 | 0.085 |
| PROMIS Questionnaires |  |  |  |  |  |
| - Pain Intensity 3a | - | - | 0.983 | 0.905 – 1.068 | 0.687 |
| - Neuropathic Pain 5a | - | - | 1.016 | 0.937 – 1.103 | 0.697 |
| - Anxiety 4a | - | - | 1.038 | 0.976 – 1.104 | 0.232 |
| - Depression 4a | - | - | 1.058 | 0.983 – 1.138 | 0.134 |
| - Physical Function 4a | - | - | 0.976 | 0.896 – 1.063 | 0.576 |
| - Sleep Disturbance 4a | - | - | 0.996 | 0.931 – 1.066 | 0.907 |
| McGill Pain Questionnaire (Score) |  |  |  |  |  |
| - Sensory component | - | - | 1.043 | 0.955 – 1.139 | 0.345 |
| - Affective component | - | - | 1.157 | 0.920 – 1.455 | 0.212 |
| - Total | - | - | 1.042 | 0.970 – 1.118 | 0.259 |
| Pain Catastrophizing Scale (Score) |  |  |  |  |  |
| - Rumination | - | - | 1.016 | 0.891 – 1.158 | 0.813 |
| - Magnification | - | - | 1.045 | 0.815 – 1.341 | 0.727 |
| - Helplessness | - | - | 1.035 | 0.913 – 1.174 | 0.592 |
| - Total | - | - | 1.012 | 0.957 – 1.070 | 0.677 |
| CI at Baseline<sup>3</sup> | 17 (21) | 6 (35.3) | 2.053 | 0.664 – 6.353 | 0.212 |

<sup>1</sup> Data are presented as 95% Confidence Interval (CI) of Odds Ratios (OR)
<sup>2</sup> Variables were selected based on previously in literature described risk factors for postoperative cognitive dysfunction (POCD), the pain, mood and sleep questionnaires, alongside with data from the baseline characteristics demonstrating a trend towards significance.
<sup>3</sup> Cognitive Impairment (CI) at baseline was defined as having a MoCA Blind score <18

## 4 DISCUSSION

This study demonstrates that the T-Cog is a feasible and effective tool for perioperative and long-term cognitive assessment, with 3-month retention rates matching the higher end of in-person studies ^9, 18^. At 3 months post-surgery, 17% of patients showed objective decline in cognitive function.

Recommendations for nomenclature for perioperative cognitive change have been proposed. Historically POCD, defined as an objective postoperative cognitive decline, has been used in many prior studies ^1^. We defined objective decline in cognitive function as a decline of ≥1 standard deviation in normative based Z-scores— relative to individual baselines— observed postoperatively on at least two cognitive tests within the same domain ^1, 6, 9, 16, 23^. The T-Cog approach, capturing multiple cognitive dimensions, yielded a POCD incidence of 17%, consistent with previous reports on POCD in similar cohorts ^2–11^. Updated recommendations propose aligning nomenclature with DSM-5 criteria for NCD, which require objective decline, SCC reported by the individual, a knowledgeable informant or the clinician and evaluation of functional ability. In our cohort, of those who reported on SCC, only 6% (5/81) met both objective decline and SCC criteria; 9% (7/81) had objective decline without SCC, and 24% (24/81) had SCC without measurable objective decline. Similar to Deiner et al. (2019), we found discrepancies between objective impairment and SCC reported by the individual only ^33^, suggesting that strict adherence to current NCD criteria may under-identify patients with meaningful objective cognitive decline. These discrepancies may reflect not only anosognosia (i.e. reduced insight due to cognitive dysfunction), but also a lack of awareness related to the subtlety of early deficits or the postoperative context, where patients may attribute changes to aging, pain, or recovery rather than cognition. This may be particularly relevant in our study, as detailed cognitive assessments across multiple domains can detect subtle but clinically significant cognitive changes that may not be readily obvious to patients themselves.

While brief tools like the telephone MoCA (T-MoCA) have shown validity for preoperative cognitive screening ^19, 20, 34^, our study is among the first to utilize a more comprehensive remote assessment approach in the perioperative period. Patients who developed objective cognitive decline, in addition to lower MoCA scores, also showed lower preoperative scores in attention, executive function, processing speed, and language domains, indicating subtle multidomain deficits not captured by brief screening instruments ^6, 35^. As perioperative NCD adversely affect recovery and increase morbidity and mortality, these findings highlight the potential value of a more comprehensive remote test battery, such as the T-cog, in improving preoperative risk stratification and facilitating postoperative cognitive monitoring, especially in older adults, and particularly those with limited post-surgical mobility ^18, 36^.

Although chronic pain has been associated with MCI in cross-sectional studies ^15^, prospective studies are needed to examine pain as an independent risk factor for cognitive decline ^2, 37^. Although our study did not yield any conclusive associations between pre-surgical pain and mood disturbances and NCD, as prior studies have done ^38, 39^, or between postoperative pain phenotypes and NCD ^40, 41^, it is likely due to the relatively modest sample size. Importantly, however, our results here demonstrate the feasibility to combine comprehensive cognitive assessment with comprehensive evaluations of mood and pain, regardless of the types of surgery (hip vs. knee arthroplasty). This technical feasibility thus enables future studies of larger scales to test these important associations.

Interestingly, we found no direct correlation between MCI and objective cognitive decline, again possibly due to sample size limitations, despite significant baseline impairments in the decline group. With 20–40% of MCI patients progressing to dementia annually ^14^, understanding its relationship to objective cognitive decline is critical. Given the long-term personal and socioeconomic consequences of objective cognitive decline, developing targeted risk mitigation strategies—including rehabilitation and intervention programs—will be important for improving perioperative outcomes ^7, 16, 17^.

Finally, our study showed that despite its length, the T-Cog is feasible for both perioperative and long-term use. A 2016 qualitative systematic review by Paredes et al. found that among 24 studies evaluating objective decline in cognitive function after non-cardiac surgery, retention rates typically ranged from 73% to 95% at three months ^8, 9^. More recent studies focused specifically on older orthopedic patients continue to report similar retention rates, with approximately 86% maintained at three months ^3^. Our follow up rate is comparable to these previous studies. Given its feasibility and comprehensive nature of assessment, it can be combined with other approaches such as electroencephalographic recordings or blood and cerebrospinal fluid analysis to enable further mechanistic or biomarker studies^42, 43^. It can also be used in clinical trials to assess therapies to treat postoperative cognitive decline, to augment studies that have focused on acute-phase postoperative delirium^44^.

This study has several limitations. As an ongoing study, the current 3-month follow-up sample size is modest; larger cohorts are thus needed to reliably test associations between NCDs, preoperative cognitive function, and preoperative pain, allowing for multivariate analyses. The absence of a control group also limits comparison, although clinically meaningful cognitive decline over such a short interval is unlikely to reflect normal aging alone. Repeated administration of the same cognitive assessment introduces the potential for recall bias, which may be more likely to happen with frequent testing. In this study, we selected multiple time points due to their relevance for PND research, and we tested the feasibility and compliance with the T-cog across all these times. Having established feasibility across multiple time points, future studies can choose time points of cognitive testing judiciously based on specific study questions to avoid recall bias. This bias can also be studied using a control cohort. It would also be interesting to compare the recall bias between shorter tests currently in use and other longer more comprehensive battery. Our patient population is old, frail, and often with multiple comorbid medical conditions, and thus there is a possibility that some of them may have experienced time-related decline in cognitive function. Thus, future studies should include control subjects with similar joint pathology who decline surgery to account for potential learning effects or natural variability in cognitive scores over time. This is a single-center study with a highly educated population, and so there is a further need to demonstrate the feasibility of T-cog as well as to quantify the incidence of cognitive decline in a larger, more heterogenous population. Overall, 18 patients were lost to follow up from the time of surgery to the third postoperative month. It is possible that a number of these patients also experienced cognitive difficulties in the postoperative period which contributed to their loss of follow up. If so, the true incidence of postoperative cognitive decline in this population may be higher than what we observed, suggesting that our estimate could be conservative. Future studies should thus complement T-cog with subjective cognitive assessment from informant or clinician on impairments in activities of daily living, which constitute criteria proposed for the diagnosis of NCD. Additionally, we have only used the English version of the T-cog, but language barrier could impact the testing of cognitive changes. Thus, in future studies, other language versions of T-cog should be applied whenever appropriate. Lastly, improving the validity of remote testing by better controlling the testing environment warrants further investigation.

In summary, at 3-month follow-up, using the remote UDS v3.0 T-cog, 17% of patients over the age 65 had objective decline in cognitive function, demonstrating testing feasibility. Patients who developed objective decline in cognitive function also had lower baseline scores in attention, executive function, processing speed and language. To our knowledge, this is the first study to integrate a comprehensive remote cognitive assessment tool with preoperative pain and mood evaluation in an older orthopedic surgical cohort, enabling future studies on long-term cognitive and pain outcomes in the perioperative setting.

## Data Availability

All data produced in the present study are available upon reasonable request to the authors.

## ACKNOWLEDGEMENTS

This study is supported by the Aging Research Community (ARCO) Initiative and the Interdisciplinary Pain Research Program (IPRP) at the New York University Grossman School of Medicine. This study was also supported by NIH grant P30AG066512 (to Dr. Thomas Wisniewski).

## CONSENT STATEMENT

The study protocol was approved by the New York University Grossman School of Medicine Institutional Review Board (6/27/2023, #123-00664). Study procedures were conducted in accordance with the ethical standards as laid down in the Declaration of Helsinki. All participants gave written informed consent.

## SUPPLEMENTARY DIGITAL CONTENT

**Figure S1. Cognitive Domains Assayed by Individual Tests**

**Figure S2. Pie Charts Illustrating the Results of the Patient Satisfaction Survey**

**Table S1. Baseline Characteristics and Outcomes According to Type of Surgery**

**Table S2. Subgroup Analysis: Cognitive Decline at 3 Months**

## REFERENCES

1. Evered L, Silbert B, Knopman DS, et al. Recommendations for the nomenclature of cognitive change associated with anaesthesia and surgery—2018. J Alzheimers Dis. 2018;66(1):1–10.

2. Huai X, Jiao Y, Gu X, et al. Preoperative Chronic Pain as a Risk Factor for Early Postoperative Cognitive Dysfunction in Elderly Patients Undergoing Hip Joint Replacement Surgery: A Prospective Observational Cohort Study. Front Neurosci. 2021;15:747362. doi:10.3389/fnins.2021.747362

3. Amirpour A, Bergman L, Markovic G, Liander K, Nilsson U, Eckerblad J. Understanding neurocognitive recovery in older adults after total hip arthroplasty—neurocognitive assessment, blood biomarkers and patient experiences: a mixed-methods study. BMJ open. 2025;15(1):e093872.

4. Atkins KJ, Silbert B, Scott DA, Evered LA. Prevalence of neurocognitive disorders 5 years after elective orthopaedic surgery. Anaesthesia. 2024;79(10):1053–1061.

5. Evered L, Scott DA, Silbert B, Maruff P. Postoperative Cognitive Dysfunction Is Independent of Type of Surgery and Anesthetic. Anesth Analg. 2011;112(5)

6. Borchers F, Spies CD, Feinkohl I, et al. Methodology of measuring postoperative cognitive dysfunction: a systematic review. Br J Anaesth. 2021;126(6):1119–1127.

7. Dilmen OK, Meco BC, Evered LA, Radtke FM. Postoperative neurocognitive disorders: a clinical guide. J Clin Anesth. 2024;92:111320.

8. Monk Terri G, Weldon B C, Garvan Cyndi W, et al. Predictors of Cognitive Dysfunction after Major Noncardiac Surgery. Anesthesiology. 2008;108(1)

9. Paredes S, Cortínez L, Contreras V, Silbert B. Post operative cognitive dysfunction at 3 months in adults after non cardiac surgery: a qualitative systematic review. Acta Anaesthesiol Scand. 2016;60(8):1043–1058.

10. Moller JT, Cluitmans P, Rasmussen LS, et al. Long-term postoperative cognitive dysfunction in the elderly: ISPOCD1 study. Lancet. 1998;351(9106):857–861.

11. Subramaniyan S, Terrando N. Neuroinflammation and perioperative neurocognitive disorders. Anesth Analg. 2019;128(4):781–788.

12. Evered L, Scott DA, Silbert B. Cognitive decline associated with anesthesia and surgery in the elderly: does this contribute to dementia prevalence? Curr Opin Psychiatry. 2017;30(3):220–226.

13. Rudolph JL, Marcantonio ER. Postoperative delirium: acute change with long-term implications. Anesth Analg. 2011;112(5):1202–1211.

14. Skolariki K, Terrera GM, Danso SO. Predictive models for mild cognitive impairment to Alzheimer’s disease conversion. Neural Regen Res. 2021;16(9):1766–1767.

15. Chen J, Wang X, Xu Z. The relationship between chronic pain and cognitive impairment in the elderly: a review of current evidence. J Pain Res. 2023:2309–2319.

16. Berger M, Nadler JW, Browndyke J, et al. Postoperative Cognitive Dysfunction: Minding the Gaps in Our Knowledge of a Common Postoperative Complication in the Elderly. Anesthesiol Clin. 2015;33(3):517–50.

17. Berger M, Schenning KJ, Brown CHI, Deiner SG, Whittington RA, Eckenhoff RG. Best Practices for Postoperative Brain Health: Recommendations From the Fifth International Perioperative Neurotoxicity Working Group. Anesth Analg. 2018;127(6):1406–1413.

18. Funder KS, Steinmetz J, Rasmussen LS. Methodological Issues of Postoperative Cognitive Dysfunction Research. Semin Cardiothorac Vasc Anesth. 2010;14(2):119–122.

19. Gierzynski TF, Gregoire A, Reader JM, et al. Evaluation of the Uniform Data Set version 3 teleneuropsychological measures. J Int Neuropsychol Soc. 2024;30(2):183–193.

20. Rockholt MM, Wu RR, Zhu E, et al. Application of the Uniform Data Set version 3 tele-adapted test battery (T-cog) for remote cognitive assessment preoperatively in older adults. Front Aging Neurosci. 2024;16:1535830.

21. Gagliese L, Melzack R. Chronic pain in elderly people. Pain. 1997/03/01/ 1997;70(1):3–14.

22. Dimitriou D, Antoniadis A, Flury A, Liebhauser M, Helmy N. Total hip arthroplasty improves the quality-adjusted life years in patients who exceeded the estimated life expectancy. J Arthroplasty. 2018;33(11):3484–3489.

23. Devinney MJ, Mathew JP, Berger M. Postoperative delirium and postoperative cognitive dysfunction: Two sides of the same coin? Anesthesiology. 2018;129(3):389.

24. Rasmussen L, Johnson T, Kuipers H, et al. Does anaesthesia cause postoperative cognitive dysfunction? A randomised study of regional versus general anaesthesia in 438 elderly patients. Acta Anaesthesiol Scand. 2003;47(3):260–266.

25. Davis N, Lee M, Lin AY, et al. Postoperative Cognitive Function Following General Versus Regional Anesthesia: A Systematic Review. J Neurosurg Anesthesiol 2014;26(4)

26. Rattanabannakit C, Risacher SL, Gao S, et al. The cognitive change index as a measure of self and informant perception of cognitive decline: relation to neuropsychological tests. J Alzheimers Dis. 2016;51(4):1145–1155.

27. Cella D, Riley W, Stone A, et al. The Patient-Reported Outcomes Measurement Information System (PROMIS) developed and tested its first wave of adult self-reported health outcome item banks: 2005–2008. J Clin Epidemiol. 2010;63(11):1179–1194.

28. Melzack R. The McGill Pain Questionnaire: major properties and scoring methods. Pain. 1975;1(3):277–299.

29. Sullivan MJ, Bishop SR, Pivik J. The pain catastrophizing scale: development and validation. Psychol Assess. 1995;7(4):524.

30. Weintraub S, Besser L, Dodge HH, et al. Version 3 of the Alzheimer Disease Centers’ neuropsychological test battery in the Uniform Data Set (UDS). Alzheimer Dis Assoc Disord. 2018;32(1):10–17.

31. Stricker NH, Christianson TJ, Lundt ES, et al. Mayo normative studies: Regression-based normative data for the auditory verbal learning test for ages 30–91 years and the importance of adjusting for sex. J Int Neuropsychol Soc. 2021;27(3):211–226.

32. Mrazik M, Millis S, Drane DL. The oral trail making test: effects of age and concurrent validity. Arch Clin Neuropsychol. 2010;25(3):236–243.

33. Deiner S, Liu X, Lin HM, et al. Subjective cognitive complaints in patients undergoing major non-cardiac surgery: a prospective single centre cohort trial. Br J Anaesth. Jun 2019;122(6):742–750.

34. Abess AT, Shah NJ, Whitlock EL, et al. Brain Health Screening in Older Surgical Patients: A Multicenter, Retrospective, Observational Analysis and Survey. Anesth Analg. May 16 2025.

35. Geddes MR, O’Connell ME, Fisk JD, et al. Remote cognitive and behavioral assessment: report of the Alzheimer Society of Canada Task Force on dementia care best practices for COVID-19. Alzheimers Dement (Ams). 2020;12(1):e12111.

36. Price CC, Burt JS, Amini S, et al. The Preoperative Phases of the Perioperative Cognitive Anesthesia Network for Older Adults Electing Surgery: Results From an Observational Cohort. Anesth Analg. Mar 6 2025.

37. Gong G-L, Liu B, Wu J-X, Li J-Y, Shu B-Q, You Z-J. Postoperative cognitive dysfunction induced by different surgical methods and its risk factors. Am Surg. 2018;84(9):1531–1537.

38. Brodier E, Cibelli M. Postoperative cognitive dysfunction in clinical practice. BJA Educ. 2021;21(2):75–82.

39. Ferrari R, Vidotto G, Muzzolon C, Auriemma S, Salvador L. Neurocognitive deficit and quality of life after mitral valve repair. J Heart Valve Dis. 2014;23(1):72–78.

40. Suraarunsumrit P, Srinonprasert V, Kongmalai T, et al. Outcomes associated with postoperative cognitive dysfunction: a systematic review and meta-analysis. Age Ageing. 2024;53(7):afae160.

41. Wang Y, Sands LP, Vaurio L, Mullen EA, Leung JM. The Effects of Postoperative Pain and Its Management on Postoperative Cognitive Dysfunction. Am J Geriatr Psychiatry. 2007/01/01/ 2007;15(1):50–59.

42. Bruzzone MJ, Chapin B, Walker J, et al. Electroencephalographic Measures of Delirium in the Perioperative Setting: A Systematic Review. Anesth Analg. May 1 2025;140(5):1127–1139.

43. Sim MA, Wilding H, Atkins KJ, Silbert B, Scott DA, Evered LA. Preoperative Biofluid Biomarkers for Predicting Postoperative Neurocognitive Disorders in Older Adults: A Systematic Review. Anesth Analg. 2025;141(3):570–587.

44. Karageorgos V, Darivianaki P, Spartinou A, et al. A Randomized Clinical Trial of Dexmedetomidine on Delirium, Cognitive Dysfunction, and Sleep After Non-Ambulatory Orthopedic Surgery With Regional Anesthesia. Anesth Analg. May 22 2025

