## Supplemental Figure 1 for "A Remote Comprehensive Neurocognitive Test Battery to Monitor Postoperative Neurocognitive Dysfunction in Older Adults: A Prospective Observational Study"

**Supplemental Figure 1.** Cognitive domains assayed by individual tests.

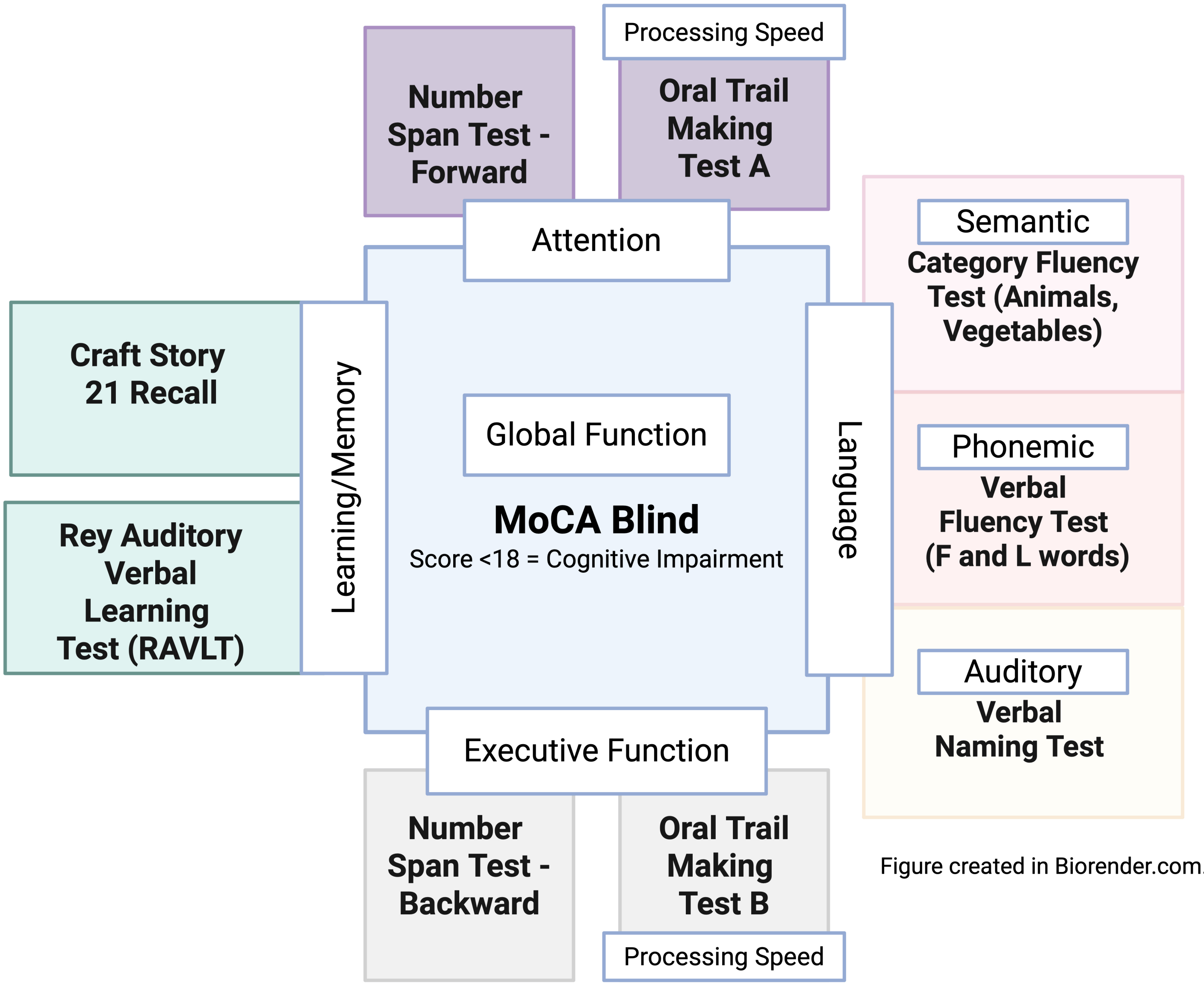


Figure created in Biorender.com.
