## Supplemental Table 1 for "A Remote Comprehensive Neurocognitive Test Battery to Monitor Postoperative Neurocognitive Dysfunction in Older Adults: A Prospective Observational Study"

**Supplemental Table 1.** Baseline Characteristics and Outcomes according to Type of Surgery.

|  | **Type of Surgery** | |  |
| --- | --- | --- | --- |
| **Demographics** | **Total Knee Arthroplasty**  **(n = 55)** | **Total Hip Arthroplasty**  **(n = 43)** | **p-value** |
| **Gender, (%)**   - *Male* - *Female* | 23 (41.8)  32 (58.2) | 19 (44.2)  24 (55.8) | 0.814 |
| **Age, mean (± SD)** | 74.0 ± 4.4 | 73.4 ± 6.1 | 0.447 |
| **Years of Education, mean (± SD)^[1]^** | 17.3 ± 3.1 | 18.0 ± 2.9 | 0.277 |
| **Race (%)^[2]^**   - *White* - *Black/African American* - *Asian* - *American Indian/Alaskan American* - *Unknown*   **Ethnicity (%)**   - *Hispanic or Latino* - *Not Hispanic or Latino* - *Not Reported/Unknown* | 41 (74.5)  8 (14.5)  2 (3.6)  0 (0.0)  4 (7.3)  8 (14.5)  38 (69.1)  9 (16.4) | 34 (79.1)  7 (16.3)  1 (2.3)  0 (0.0)  1 (2.3)  2 (4.7)  39 (90.7)  2 (4.6) | 0.979  0.090 |
| **Past Medical History, yes (%)**   - *Hypertension* - *Hypercholesterolemia and/or hyperlipidemia* - *Obesity* - *History of Malignancy (Active or Past)* - *Thyroid Disease* - *Anxiety* - *Depression* - *Lung Disease (Asthma and/or COPD)* - *Coronary Artery Disease* - *Diabetes* - *Sleep Apnea*   **Smoking History (Current or Former), yes (%)**  **History of Alcohol Overconsumption, yes (%)**  **Medication History (%)**   - Opioids - Anti-inflammatory Agents (NSAIDs) - Gabapentinoids - Benzodiazepines - Antidepressants | 37 (67.3)  39 (70.9)  24 (43.6)  18 (32.7)  14 (25.5)  7 (12.7)  12 (21.8)  3 (5.5)  10 (18.2)  12 (21.8)  11 (20.0)  31 (56.4)  8 (14.5)  18 (32.7)  28 (50.9)  4 (7.3)  6 (10.9)  9 (16.4) | 29 (67.4)  26 (60.5)  12 (27.9)  13 (30.2)  8 (18.6)  5 (11.6)  4 (9.3)  7 (16.3)  7 (16.3)  8 (18.6)  6 (14.0)  17 (39.5)  6 (14.0)  12 (27.9)  23 (53.3)  7 (16.3)  3 (7.0)  6 (14.0) | 0.986  0.278  0.109  0.792  0.420  0.869  0.096  0.079  0.805  0.695  0.433  0.098  0.934  0.607  0.800  0.161  0.504  0.742 |
| **Type of Anesthesia, (%)**   - *Regional Anesthesia* - *General Anesthesia* | 52 (94.5)  3 (5.5) | 41 (95.3)  2 (4.7) | 0.858 |
| **Cognitive Outcomes^[3]^** | | | |
| **Post-surgical Objective Cognitive Decline, yes (%)**   - At 1 week - At 1 month - At 3 months   **Subjective Cognitive Complaints, yes (%)**   - *Missing Data* | 9 (18.4)  10 (19.6)  11 (20.0)  17 (39.5)  12 (21.8) | 4 (10.8)  3 (7.3)  6 (14.0)  11 (29.7)  6 (14.0) | 0.333  0.093  0.433  0.359 |
| **Pain, Mood, and Sleep Questionnaires: Baseline Scores^[4]^** | | | |
| **PROMIS Questionnaires (T-Score)**   - *Pain Intensity 3a* - *Neuropathic Pain 5a* - *Anxiety 4a* - *Depression 4a* - *Physical Function 4a* - *Sleep Disturbance 4a*   **McGill Pain Questionnaire (Score)**   - *Sensory component* - *Affective component* - *Total*   **Pain Catastrophizing Scale (Score)**   - *Rumination* - *Magnification* - *Helplessness* - *Total* | 52.5 ± 6.9  44.9 ± 7.6  50.0 ± 9.7  47.1 ± 7.4  37.3 ± 7.2  49.4 ± 8.7  11.7 ± 6.8  2.6 ± 2.4  14.3 ± 8.4  3.8 ± 4.5  2.0 ± 2.6  3.9 ± 4.4  9.6 ± 10.4 | 51.9 ± 6.6  45.1 ± 6.2  50.9 ± 8.7  47.3 ± 7.4  36.8 ± 7.0  53.3 ± 8.5  10.9 ± 6.0  2.6 ± 2.4  13.6 ± 7.5  3.9 ± 4.3  1.6 ± 1.8  3.4 ± 4.5  8.9 ± 9.6 | 0.510  0.842  0.592  0.910  0.622  0.045  0.598  0.976  0.675  0.942  0.729  0.433  0.623 |
| **Pain, Mood, and Sleep Questionnaires: 3-months^[5]^** | | | |
| **PROMIS Questionnaires (T-Score)**   - *Pain Intensity 3a* - *Neuropathic Pain 5a* - *Anxiety 4a* - *Depression 4a* - *Physical Function 4a* - *Sleep Disturbance 4a*   **McGill Pain Questionnaire (Score)**   - *Sensory component* - *Affective component* - *Total*   **Pain Catastrophizing Scale (Score)**   - *Rumination* - *Magnification* - *Helplessness* - *Total* | 46.8 ± 7.0 (↓)  45.6 ± 6.6 (↑)  46.5 ± 8.3 (↓)  45.7 ± 6.9 (↓)  41.3 ± 7.2 (↑)  49.1 ± 9.2 (↓)  7.3 ± 6.7 (↓)  1.4 ± 1.7 (↓)  8.6 ± 8.0 (↓)  2.1 ± 2.1 (↓)  0.8 ± 1.3 (↓)  2.4 ± 3.2 (↓)  5.3 ± 5.7 (↓) | 39.1 ± 7.7 (↓)  40.0 ± 4.6 (↓)  46.2 ± 8.0 (↓)  44.5 ± 5.8 (↓)  43.3 ± 9.6 (↑)  47.1 ± 10.3 (↓)  2.9 ± 3.7 (↓)  0.7 ± 1.2 (↓)  3.6 ± 4.6 (↓)  1.0 ± 1.5 (↓)  0.8 ± 1.3 (↓)  1.0 ± 1.3 (↓)  2.8 ± 3.2 (↓) | <0.001  <0.001  0.860  0.442  0.370  0.354  <0.001  0.089  <0.001  0.012  0.942  0.093  0.039 |

1. Years of education was calculated as the summarized amount of years according to previously plotted neuropsychiatric data, with finished: high school = 12 years, bachelors = 4 years, masters = 2 years, doctoral = 4 years, associate degree = 2 years (only if completed).
2. Because some cells had expected counts<5, we used Fisher’s exact test to compare distributions of Race and Ethnicity between knee and hip arthroplasty groups.
3. Objective cognitive decline was defined as a decline of ≥1 standard deviation in normative based Z-scores—relative to the individual's own baseline—observed at the postoperative time point on at least two cognitive tests in the UDS v3.0 T-cog test battery within the same domain. Subjective Cognitive Complaints (SCC) was evaluated using an open-ended question (yes with explanation in free text/no) and/or the 20-item cognitive change index (CCI-20).
4. Scores are mean ± SD.
5. Arrows in brackets indicate changes in performance relative to baseline (↑ denotes improvement, ↓ denotes deterioration), except for the PROMIS Physical Function 4a, where increases in T-score signify an improvement.
