## Supplemental Table 2 for "A Remote Comprehensive Neurocognitive Test Battery to Monitor Postoperative Neurocognitive Dysfunction in Older Adults: A Prospective Observational Study"

| **Subgroup analysis: Cognitive Decline at 3 months (n = 17)^[1]^** | | | | | |
| --- | --- | --- | --- | --- | --- |
| **Subject** | **CI at**  **Baseline^[2]^** | **1-week^[3]^** | **1-month^3^** | **3-months^3^** | **SCC^[4]^** |
| 1 | No | Yes (Executive Function/Processing Speed) | Yes (Executive Function/Processing Speed) | Yes (Executive Function/Processing Speed) | No |
| 2 | No | Yes (Executive Function/Processing Speed) | Yes (Executive Function/Processing Speed) | Yes (Executive Function/Processing Speed) | - |
| 3 | No | Yes (Learning/Memory) | Yes (Memory and Executive Function) | Yes (Executive Function/Processing Speed) | - |
| 4 | No | No | Yes (Learning/Memory) | Yes (Learning/Memory) | - |
| 5 | No | Yes (Learning/Memory) | No | Yes (Learning/Memory) | Yes |
| 6 | Yes | Yes (Executive Function/Processing Speed) | Yes (Executive Function/Processing Speed) | Yes (Executive Function/Processing Speed) | No |
| 7 | No | No | Yes (Attention/Executive Function) | Yes (Attention/Executive Function) | Yes |
| 8 | Yes | - | - | Yes (Executive Function/Processing Speed) | Yes |
| 9 | No | No | - | Yes (Attention/Processing Speed) | No |
| 10 | No | No | No | Yes (Executive Function/Processing Speed) | No |
| 11 | Yes | - | Yes (Memory/Executive Function) | Yes (Learning/Memory)^[5]^ | - |
| 12 | No | - | - | Yes (Memory/Language)^[6]^ | Yes |
| 13 | Yes | No | Yes (Learning/Memory)^[7]^ | Yes (Learning/Memory) | No |
| 14 | No | - | - | Yes (Learning/Memory)^6^ | - |
| 15 | Yes | - | No | Yes (Learning/Memory)^7^ | No |
| 16 | No | No | - | Yes (Learning/Memory) | Yes |
| 17 | Yes | - | No | Yes (Attention/Processing Speed) | No |

**Supplemental Table 2.** Subgroup analysis for patients who developed cognitive decline after surgery.

1. Objective cognitive decline was defined as a decline of ≥1 standard deviation in Z-scores—relative to both the group baseline and the individual's own baseline—at each postoperative time point, in at least two cognitive tests within the same domain, as measured using the UDS v3.0 T-cog test battery. The specific cognitive domains assessed for each subject are indicated in parentheses.
2. Cognitive Impairment (CI) at baseline was defined as having a MoCA blind score <18.
3. All examinations rated as “Very Valid” are highlighted in green, those rated as “Questionably Valid” in yellow, and those rated as “Invalid” in red. Exams are left white if the validity was not assessed Missed exams were marked as “-“.
4. Subjective Cognitive Complaints (SCC). Empty answered (no response) were marked as “-“.
5. Reason for questionably valid exam: potential language barrier
6. Reason for questionably valid exam: distractions

   ^7^ Reason for questionably valid exam: hearing impairment and distractions.
