## Supplementary figures and images for "A Remote Comprehensive Neurocognitive Test Battery to Monitor Postoperative Neurocognitive Dysfunction in Older Adults: A Prospective Observational Study"

### Supplemental Figure 2

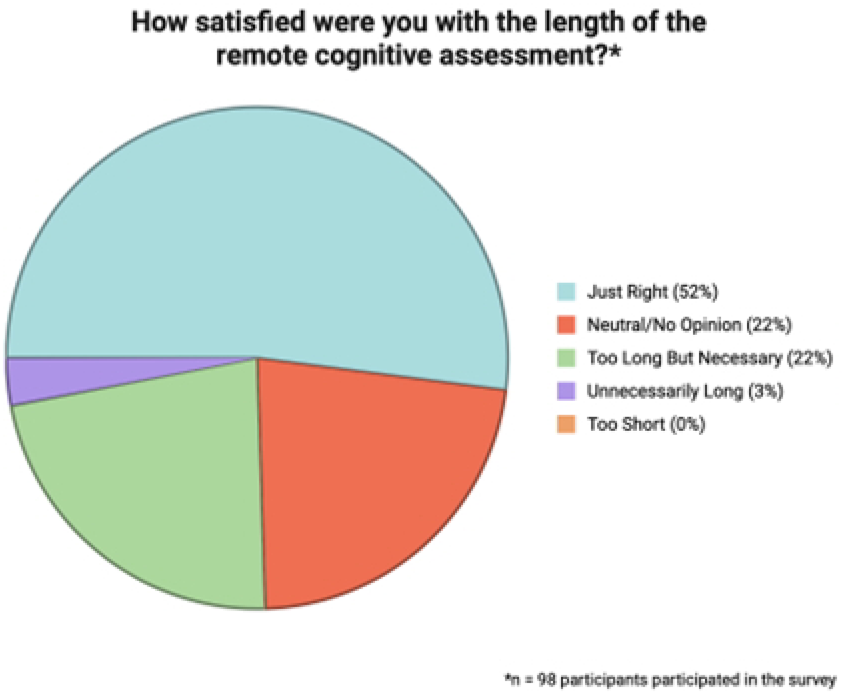

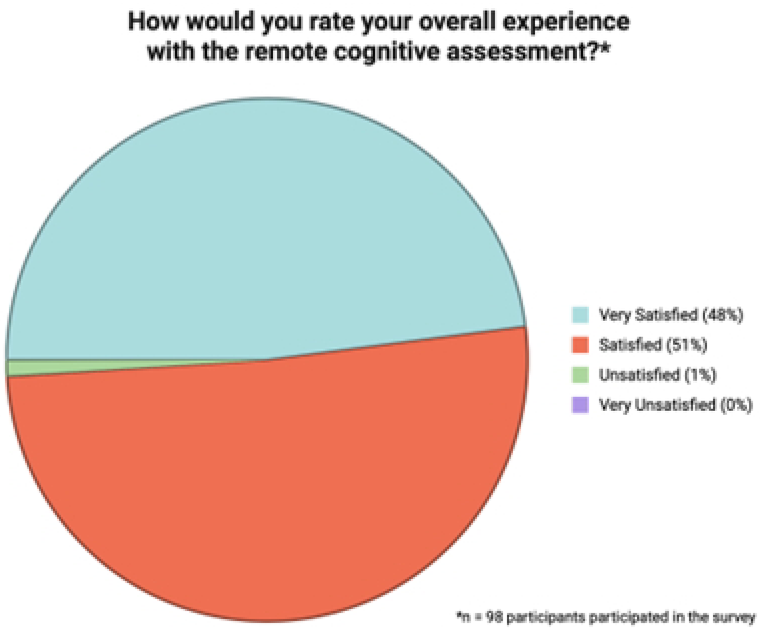

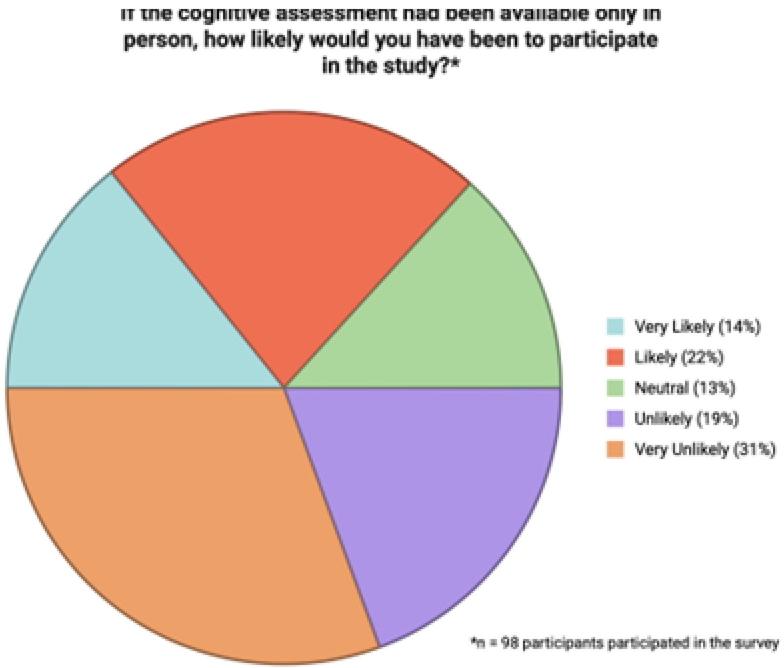

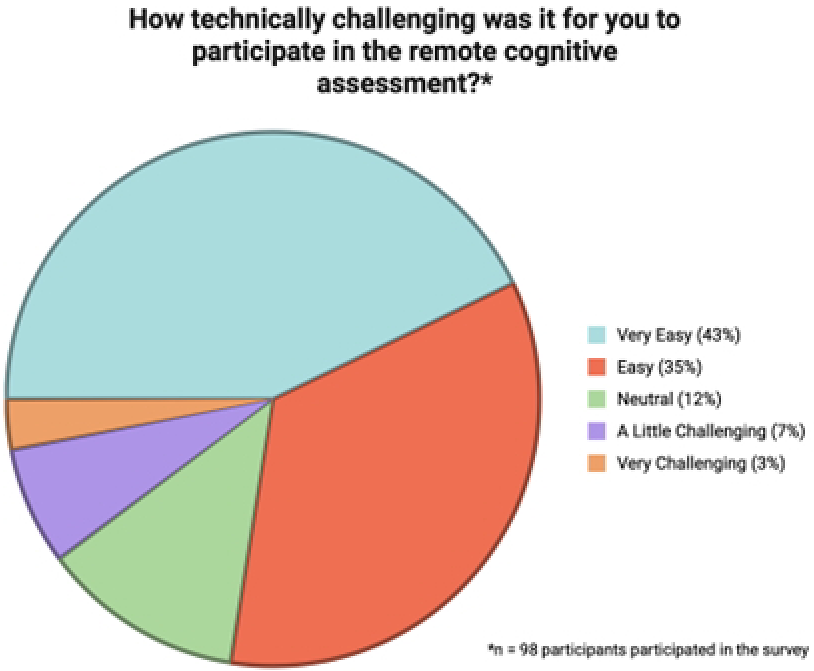
